# Effect of COVID-19 response policies on walking behavior in US cities

**DOI:** 10.1101/2020.12.07.20245282

**Authors:** Ruth F. Hunter, Leandro Garcia, Thiago Herick de Sa, Belen Zapata-Diomedi, Christopher Millet, James Woodcock, Alex ’Sandy’ Pentland, Esteban Moro

## Abstract

The COVID-19 pandemic has caused mass disruption to our daily lives. Mobility restrictions implemented to reduce the spread of COVID-19 have impacted walking behavior, but the magnitude and spatio-temporal aspects of these changes have yet to be explored. Walking is the most common form of physical activity and non-motorized transport, and so has an important role in our health and economy. Understanding how COVID-19 response measures have affected walking behavior of populations and distinct subgroups is paramount to help devise strategies to prevent the potential health and societal impacts of declining walking levels. In this study, we integrated mobility data from mobile devices and area-level data to study the walking patterns of 1.62 million anonymous users in 10 metropolitan areas in the United States (US). The data covers the period from mid-February 2020 (pre-lockdown) to late June 2020 (easing of lockdown restrictions). We detected when users were walking, measured distance walked and time of the walk, and classified each walk as recreational or utilitarian. Our results revealed dramatic declines in walking, especially utilitarian walking, while recreational walking has recovered and even surpassed the levels before the pandemic. However, our findings demonstrated important social patterns, widening existing inequalities in walking behavior across socio-demographic groups. COVID-19 response measures had a larger impact on walking behavior for those from low-income areas, of low education, and high use of public transportation. Provision of equal opportunities to support walking could be key to opening up our society and the economy.

## Introduction

In response to the COVID-19 outbreak, many countries have implemented interventions to induce mobility restrictions and force their citizens to stay at home (i.e., confinement) to reduce the transmission rate and prevent health services being overwhelmed. As a result, at time-points during the pandemic, over half of the world’s population have been committed to stay at home for different periods of time, causing major disruptions to their daily lives. As time has elapsed and countries are learning how to live with COVID-19, countries ebb and flow out of confinement and other social distancing policies, trying to maintain a difficult balance between reducing the transmission rate of the virus and preserving a functioning economy. Although key to containing the spread of the virus, such restrictions have caused large-scale disruption to our normal lives and active living. Preliminary descriptive studies have shown a large decrease in country-level physical activity because of the stay-at-home recommendations and strict lockdown measures^1^. Even before the COVID-19 pandemic, physical inactivity was highly prevalent – 27.5% of adults and 80% of adolescents worldwide did not meet the recommended levels for health benefits^2^,^3^ – and the fourth risk factor contributing 6% to global mortality (WHO, 2014). If we assume at least half of the world’s population have some level of lockdown restrictions in place due to the COVID-19 pandemic, then physical inactivity rates could reach > 1.1 billion people^4^,^5^.

Physical activity has a range of benefits that includes physical and mental health, strengthening of social interactions and social capital, and economic and environmental returns^6^,^7^. Walking is the most popular and accessible type of physical activity behavior. For instance, in the US, walking is consistently the most prevalent leisure-time physical activity and the most frequently reported physical activity among adults who meet public health physical activity recommendations^8^. In the US, 50.0% [95% CI 49.1%-51.0%] of adults engage in leisure walking activity and around 29.4% [95% CI 28.6%-30.3%] in utilitarian (transportation, shopping, routine) walking^8^. Despite these figures, only half of Americans self-report that they meet a minimum of 30 minutes of walking five or more times per week^9^. Utilitarian walking is in general 20% shorter but more prevalent than leisure walking^10^.

There is no doubt that mobility restrictions implemented to reduce the spread of COVID-19 have impacted walking behavior, but the magnitude and spatio-temporal aspects of these changes have not been thoroughly demonstrated yet. Much less is known on the differential impacts of COVID-19 response measures on walking behavior of population subgroups. The COVID-19 pandemic is occurring in a context of social and economic inequalities in existing non-communicable disease (NCD) and the social determinants of health^11^. The impact of COVID-19 on health inequalities will be driven by both virus-related infection and mortality rates and the consequences of the policy responses undertaken. For instance, different population subgroups are likely to have distinct experiences of lockdown according to their access to green space and urbanity, with unequal effects on their physical and mental health^11^. Given the large pre-pandemic inequalities in drivers and patterns of walking behavior, it should be expected that the impact of COVID-19 response measures on walking would show large variability between socio-demographic groups. Walking activity is unequally distributed among different socio-demographic groups. Men are more likely to walk for utilitarian purposes, but less likely for leisure, than women^8^. Prevalence of utilitarian walking decreases with increasing age, whereas prevalence of leisure walking peaks among older adults. Prevalence of walking in either context tends to increase with increasing education level and with decreasing adiposity^12^. At the same time, COVID-19 pandemic and response measures had different effects across racial and economic groups^13^. Higher COVID-19 death rates were found in the most disadvantaged areas (% of persons living in poverty, % of crowded households, and concentration of extreme racial and socioeconomic segregation) and areas with the largest populations of people of color (% of population that is non-white, Non-Hispanic).

Understanding how COVID-19 response measures have affected walking behaviour of populations and its distinct subgroups is important information to help devise strategies to prevent the potential health and societal impacts of declining walking levels^1^. To do so, a deep, at-scale understanding that goes beyond mere descriptive analysis is required. Indirect impacts of the pandemic on health, including through reductions in physical activity^14^, have been described, but mainly based on small-scale, self-report surveys and qualitative methods (e.g.^15^–17), which have well-documented biases^18^ and, sometimes, limited generalizability. These studies have also tended to focus on overall physical activity levels, with little attention to date on specific activities such as walking. Furthermore, the lack of specificity in distinguishing between utilitarian and recreational walking weakens the power of previous walking behavior analyses, with subsequent implications on policy recommendations.

In this study, we analyze changes in walking behavior using mobile phone’s geolocalized mobility data from hundreds of thousands of people in urban areas. Mobile phones are a powerful tool with which to study large-scale population dynamics, revealing patterns of human movement at greater temporal and spatial granularity, while ensuring anonymity and user privacy. Smartphones with in-built accelerometers enable 24-hour automatic recording of physical activity, providing a scalable tool to measure and quantify walking behavior^19^. We integrate anonymized and privacy-enhanced data from mobile devices and area-level data to study the walking patterns of 1.62 million anonymous users in 10 metropolitan areas in the US to investigate the effect on walking of COVID-19 response measures. The data covers the period from mid-February 2020 (pre-lockdown) to late June 2020 (easing of lockdown restrictions) in the following metropolitan areas in the US: New York, Los Angeles, Chicago, Boston, Miami, Dallas, San Francisco, Seattle, Philadelphia and Washington DC. Using simple algorithms (see Materials and Methods), we detected when users were walking, measured distance walked and time of the walk, and classified each walk as recreational or utilitarian. We also investigated walking levels during different periods of the COVID-19 response measures by population subgroups according to demographic, social, health, and built environment aspects.

## Results

### Effect of measures on utilitarian and leisure walking

Although COVID-19 response measures differed between metropolitan areas, in most of the US, social distancing started after the declaration of a national emergency on March 13th 2020 and the schools and non-essential business closings during the following week. As we can see in Figure 1, that translated into a substantial decrease of approximately 75% in the number of walks (bouts) and 55% in the average distance walked in all metropolitan areas. Since mid-April 2020, walking steadily increased although even after resuming some commercial and business activities, walking was still around 17% below the pre-pandemic level. Walking decreased considerably in Miami (−33.2%), San Francisco (−26.4%), Los Angeles (−24.8%), and New York (−23.6%), whereas in Boston (+0.6%) and Chicago (+7.15%), the distance walked was even slightly higher than before the pandemic.

**Figure 1.**
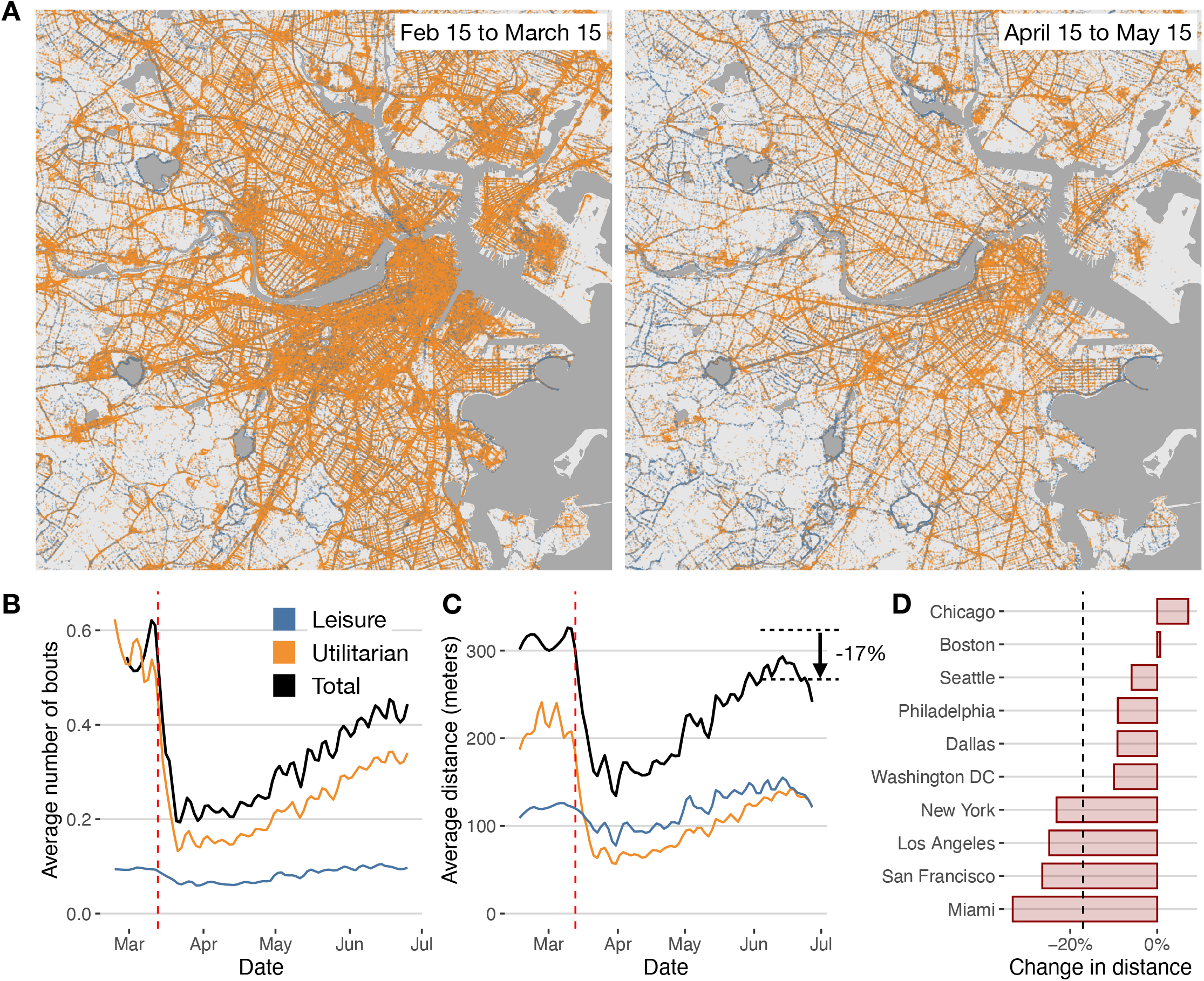
Decrease in walking behavior during the pandemic. A) Geolocations of leisure and utilitarian walks in the Boston area before (left) and after (right) the introduction of COVID-19 response measures. B) Total (black) daily average number of bouts of walking by day and user in the 10 metropolitan areas, compared with those for utilitarian and leisure walks. Vertical (red) dashed line indicates March 13th 2020, the declaration of a national emergency. C) Same as in B) but for total distance walked. D) Relative change in distance walked by city between the pre-lockdown (Feb 15 2020 to March 15 2020) and post-lockdown (June 2020).

To understand the reasons for the substantial impact of the COVID-19 response measures in walking behavior, we investigated the origin and nature of walking behavior in our cities. Our analysis showed that 87% of users undertook at least one bout of walking per week before COVID-19 response measures were introduced (March 15th 2020) and 64% walked at least 10 minutes per week. Our findings are comparable to the National Health Interview Survey (NHIS) 2010 results (self-reported 61%,^8^). On average, individuals did 0.52 bout of walking per day, similar to what was observed in the NHIS 2017 (0.49%,^20^). We found that only 4.3% of users walked at least 30 min in 5 or more days per week, comparable with the 6% reported in the NHIS^12^. Walks were typically short, with an average distance of 651.66 meters [95% CI 651.52, 651.82] and duration of 10.73 minutes [95% CI 10.73, 10.73].

To understand the nature of the walks, we used a simple classification of the bouts in utilitarian and leisure walking based on the distance walked and their final destination (see Materials and Methods). We found that most of the walks (87%) were utilitarian, but leisure walks were longer (1444.44 meters, 95% CI 1444.09, 1444.80) than utilitarian walks (407.16 meters, 95% CI 407.08, 407.24). Thus, a larger proportion of the distance walked in US metropolitan areas corresponded to leisure walking. During weekdays, most of the utilitarian walks occurred around 7am and from 3pm to 5pm, while they concentrated mostly around lunch time during the weekends (see Figure 2).

**Figure 2.**
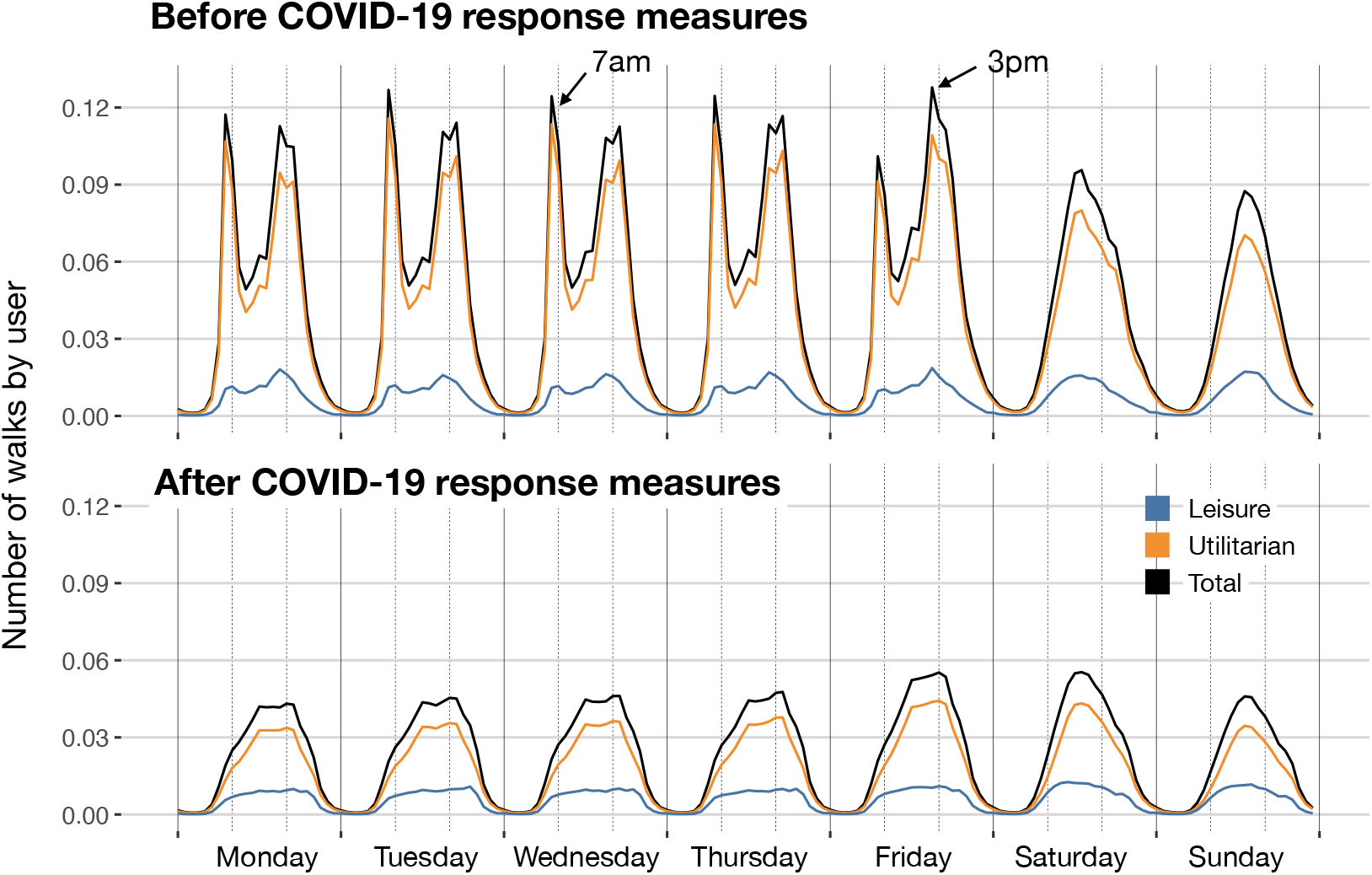
Temporal patterns of walking behavior. Panels show the average number of walks by user for each hour and day of the week, and those for leisure and utilitarian purposes. Upper panel corresponds to the temporal pattern before COVID-19 response measures and the lower panel after COVID-19 response measures.

As we can see in Figure 1, the impact of COVID-19 response measures significantly impacted utilitarian walking (−72.3% in the distance travelled directly after the declaration of a national emergency). Figure 2 shows a sharp decrease in the number of walks around 7am and 3pm, and most of the walking activity during weekdays resembled that of weekends. Our results suggest that the large impact of COVID-19 response measures in walking was mostly due to the interruption of our working, shopping and dining activities. Even after the easing of the restrictions (after mid-May 2020), utilitarian walking was still −39.2% (in distance travelled) less than before the pandemic. Leisure walking was not affected as much (Figure 1C) and recovered and surpassed the levels before the pandemic.

### Impact by socio-demographic groups

Due to the anonymous nature of our location data, we examined the socio-demographic differences at the area level, which are defined by census tracts where users live. Figure 3 shows the changes in the total number of walks and distance walked across different socio-demographic groups. Before the introduction of COVID-19 response measures, we can clearly see that groups of low income, low obesity prevalence, high park access, or high use of public transportation had higher walking activity. After the introduction of COVID-19 response measures, this was still true for obesity prevalence, park access or use of public transportation, although the relative change differed across these groups (see Figure 3C). People living in areas with high use of public transportation experienced a sharp decrease in walking activity compared to those living in areas of low use, probably due to the reduction of walking to transportation. Income groups reverse their behaviors (see Figure 3A): before COVID-19 response measures, people living in high-income areas walked less than those living in low-income areas; however, the pattern reversed after COVID-19 response measures. Furthermore, walking levels recovered to pre-COVID-19 measures in high-income areas, whereas low-income areas were still well below pre-COVID-19 levels. Figure 3B provides insights into the potential reasons for the differential observed across income groups. Utilitarian walking has decreased across all income quintiles between February 2020 and June 2020. However, people living in high-income areas have increased their leisure walking considerably (48%). This result might be related to the fact that high-income individuals had more opportunities for social distancing by staying and working at home, and also more free time and opportunities to engage in leisure walking. The substitution of utilitarian by leisure walking was not present in low-income groups and, as a result, the COVID-19 response measures have had a strong impact on their walking behavior.

**Figure 3.**
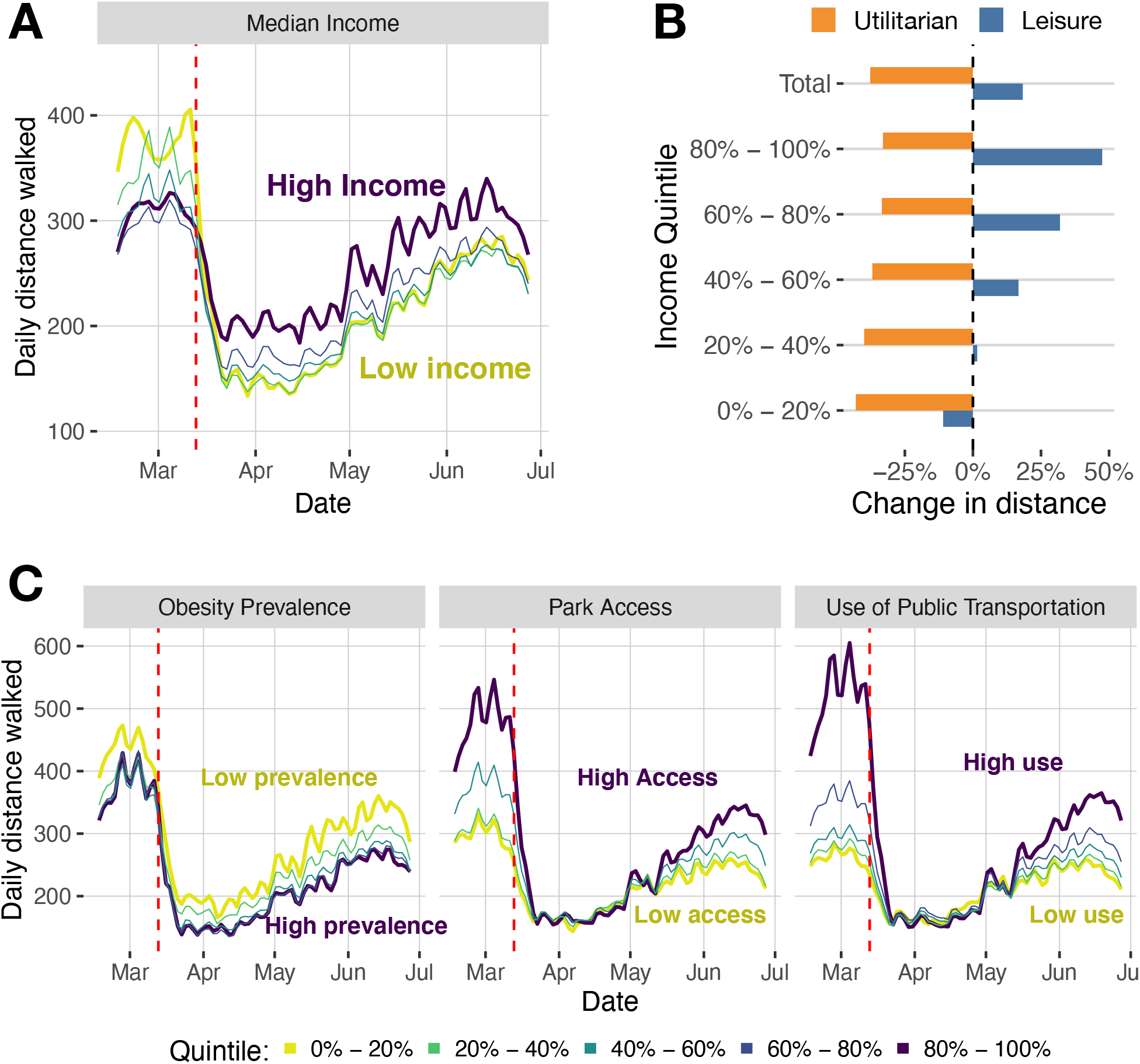
Change in walking by different socio-demographic groups. A) Average daily distance walked by different income sub-groups (quintiles). B) Change in utilitarian or leisure walking for the same income sub-groups, including the total. C) Evolution of the average distance walked by day by different sub-groups (quintiles) for obesity prevalence, park access and use of public transportation.

Using multivariate linear regression, we investigated the combined effect of area level socio-demographic (income, race, age), health (obesity prevalence), and environmental (park access) variables on the amount of walking pre-pandemic and after COVID-19 response measures were introduced. Table 1 shows the use of public transportation was the most relevant to understanding walking activity: one standard deviation in the use of public transportation increased the amount of individual walking in 61 meters by day. Areas with higher income, more black people, higher obesity prevalence or with larger fraction of 64 year-old or older walked less on average, while those living close to a park walked more. After the introduction of COVID-19 response measures, the use of public transportation was still the most impactful variable to understand walking activity. But, as noted before, the effect of the variables was different from the situation before COVID-19 response measures were introduced: areas with higher income or more park access seemed to walk more because they engaged more in leisure walking. However, areas with more black people or more obesity prevalence walked less on average.

**Table 1.**
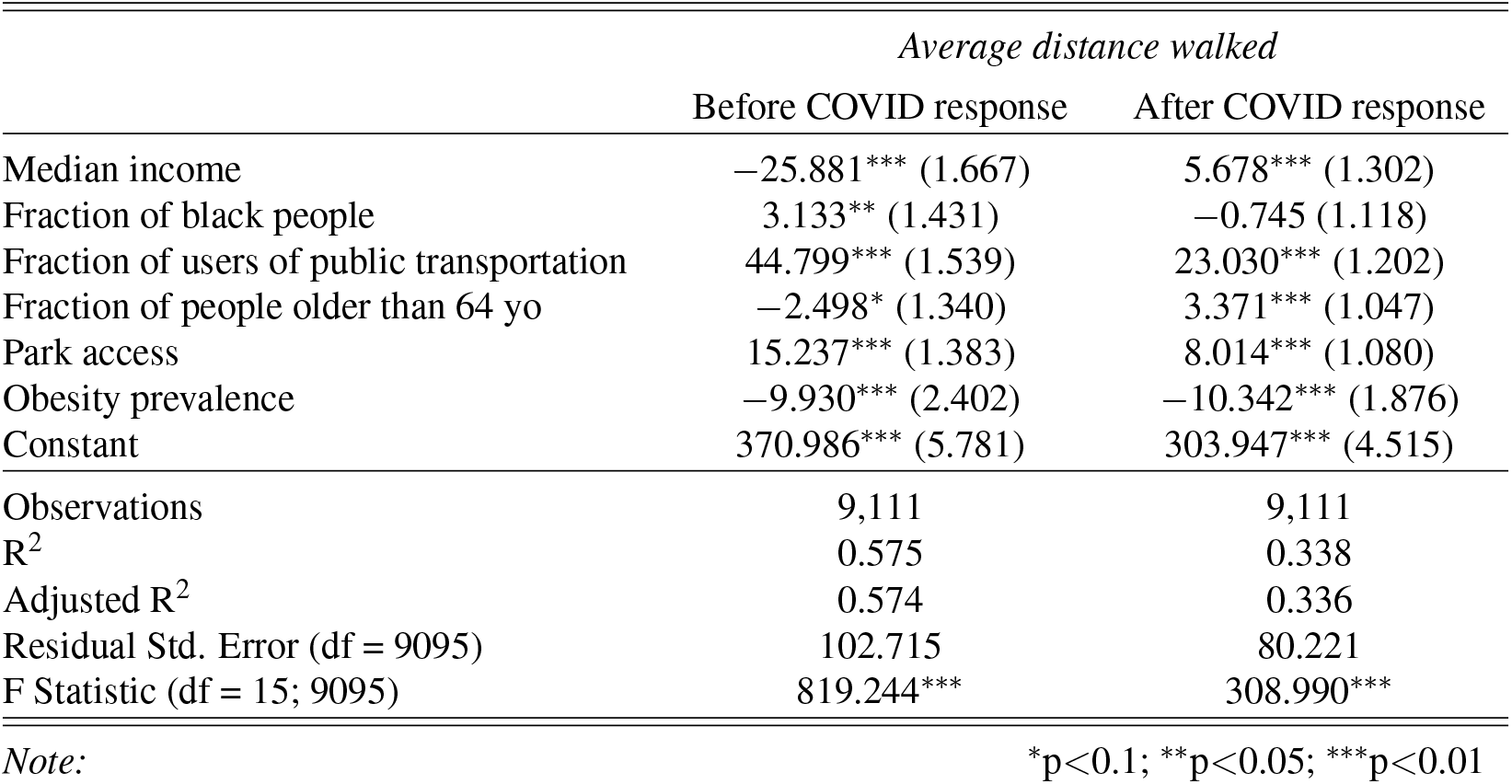
Regression results for the multivariate model (see Eq. (1) in Materials and Methods) to explain the average distance walked (in meters) by individuals in different areas as a function of the demographic properties of that area. The table shows the standardized coefficients for the regression models before and after the COVID-19 response measures were introduced. See Methods.

## Discussion

COVID-19 response measures resulted in large-scale disruption to our daily walking behavior. Our findings reveal that in all 10 metropolitan areas investigated, utilitarian walking decreased dramatically at the beginning of the lockdown restrictions owing to reductions in the needs and opportunities to walk to work, to public transport, to shop and to other amenities. The decrease in walking for leisure was less pronounced in general and, for some demographic groups, it has surpassed levels before the pandemic.

### The tale of two pandemics

Our findings also demonstrate an important differential impact on walking behavior according to demographic, social, health, and built environment factors, exacerbating existing inequalities. We show that COVID-19 response measures had a larger impact on walking behavior for those from socially disadvantaged areas, with larger fractions of public transportation users, worse obesity indicators, and a physical environment that is less conducive to walking. On the contrary, high income areas have substituted utilitarian walking activity for more leisure walking, sometimes reaching even larger levels of walking than before the pandemic.

The COVID-19 pandemic has caused disruption at scale, and requires an unprecedented level of response if we are to combat the emergent findings showing the impact on walking behavior and widening of inherent inequalities. The argument of a ‘physical inactivity pandemic’ has been made before^4^, and some have described the current COVID-physical inactivity interaction as a ‘tale of two pandemics’^21^. But COVID-19 has shown us what a political response to a ‘real’ pandemic looks like. We have witnessed determined and swift actions of governments around the world to the public health threat from COVID-19. And there is an opportunity for the same governmental determination to take radical decisions on walking behavior and tackling inherent inequalities. For example, COVID-19 response measures in Milan, Paris, and London, included pedestrianizing streets and expanding cycle lanes, facilitating COVID-19-safe transport during the crisis, while enhancing economic activity and quality of life afterwards^22^.

Provision of equal opportunities to support walking could be key to opening up our society and the economy. For example, our findings showed significant changes to utilitarian walking behavior, and with social distancing restrictions placed on local public transport, provision of environments to support utilitarian walking (and indeed cycling) provides a means of opening up the economy through return to work, and a move from home working to the office environment to support city centre economy. In the US, public transport use is typically higher in low-income communities^23^, which according to our findings suffered the largest decrease in walking behavior. Care must be taken to ensure that provision of public transport is safe, including COVID-19 mitigation strategies such as limiting the number of users, ensuring physical distancing, mandatory use of face coverings by passengers, provision of hand sanitizer, and regular cleaning of vehicles touch-points, seating etc. Albeit, the implementation of such strategies will have profound economic impacts for public transport providers.

Provision of support for leisure walking facilitates good health and well-being during social distancing and lockdown restrictions, and long-term, reduces risk of obesity and NCD, which are significant risk factors for COVID-19^14^. Supportive environments include, for example, availability of infrastructure such as access to green space (e.g. keeping public parks open) to support walking and other physical activity opportunities.

Our findings have provided evidence of the impact of COVID-19 responses on short-term inequalities for walking, and walking opportunities. If significant action is not taken, then these inequalities will likely widen in the longer term. For instance, although those residing in areas with the highest park access scores suffered the largest reductions in walking behaviour, their average daily walking distance started to build up sooner, while distance walked by those living in areas with less access to parks did not rise at the same pace. It is important that the public health community consider what adaptive and mitigation measures can be introduced to limit the effect.

### Opportunities for tactical urbanism

Some cities, especially in Europe, have sought to mitigate the impact of COVID-19 in walking behavior through temporary measures, including extending footpaths, protecting commuters using public transit through social distancing measures, pop up cycle lanes. The impact of these measures on walking behavior, especially on their disparities, warrants careful evaluation. Governments in many settings have indicated that these changes will be permanent and probably will influence walking behavior and healthy urban planning for decades. This highlights the opportunity that the COVID-19 pandemic presents for restructuring societies to meet health and environmental goals, including through transport planning. With the emergence of candidate vaccines, and substantial normalisation of societies likely by the summer, there remains a crucial opportunity to build on this momentum to make permanent changes to transport infrastructure to encourage greater walking behavior.

Our study provides valuable methodological innovation and data to quantify walking patterns and inform future city planning and policy decisions. Mobile phone data can be used to inform local policies and practice in near to real time, disentangling implications of different aspects and phases of COVID-19 response strategies for different forms of walking behavior (i.e., utilitarian and recreational) to help tailor future policies and response measures that can provide suitable walking environments to support the health and economic futures of communities.

In Table 2, we outline a range of possible low-cost interventions and policies that could increase utilitarian and recreational walking behavior and tackle the inequalities highlighted above^24^. Some examples include creating ‘pop-up’ footpaths and widening pavements which provide more opportunities to walk. Reducing speed limits in urban areas to promote pedestrian safety, and simple adaptations to adjust the timing of traffic lights to favor pedestrians provide low-cost solutions to support walking. In the long term, if made permanent, these interventions also provide an opportunity to improve population health and well-being. Other co-benefits may also include reduced air and noise pollution, reduced risk of traffic collisions and casualties, reduced risk of NCD, promoting social equity and reducing the demand on our health services. We, therefore, have a unique opportunity to improve our urban environments to support and enable walking and physical activity, particularly for our most vulnerable communities.

**Table 2.** Examples of tactical urbanism strategies that can lead to permanent solutions to increase utilitarian and leisure walking behavior and reduce inequalities. Adapted from^24^ and^22^

|  |
| --- |
| **1. Expanding pedestrian spaces and footpath networks**<br>• Programmes to encourage walking<br>• Widen and lengthen pavements and improve connections to promote traffic safety<br>• Reduce pressure on parks and public spaces by widening pavements in the surrounding area<br>• Facilitate access to vulnerable groups who would otherwise have to walk in worse environments (e.g. unsafe streets; crowded footpaths)<br>• Expand pedestrian infrastructure using existing proposals<br>• Expand pedestrian access to the provision of safe public transport |
| **2. Providing recreational paths for walking**<br>• Create and expand existing networks<br>• Establish social networks to enable and support walking |
| **3. Adapting parks and public spaces**<br>• Keep open large public spaces (e.g. parks)<br>• Ensure that parks and public space users have access to water, hygiene and sanitation.<br>• Implement and enforce measures to increase perceived and objective safety against violence, harassment. |
| **4. Adapting traffic lights, signage and speed limits**<br>• Replace button-activated traffic lights with automatic systems.<br>• Adjust the timing of traffic lights to favor pedestrians<br>• Reduce speed limits throughout the city to promote pedestrian safety<br>• Implement and enforce measures to increase pedestrian safety on traffic |

### Strengths and limitations

We investigated the impact of the COVID-19 pandemic on walking behavior in 10 of the largest US metropolitan areas using high-resolution mobility data. This data encompasses more than 1.6 million people and a wide variation in socio-demographic, health, and built environment aspects. The US is an important exemplar internationally due to having the highest number of COVID-19 cases globally, its variations across metropolitan areas in existing active travel infrastructure and pre-pandemic walking behavior, and stark inequalities in walking behavior and health outcomes that should be explicitly considered within the COVID-19 response.

Walking is the most common physical activity, but our dataset may fail to capture time spent walking when users did not carry their phone, and systematic differences may exist in wear time based on individual factors such as gender and age. Further, if we had demographic data associated with individuals’ mobility data, we could directly examine walking behavior across demographics at the individual level. However, the anonymous location data does not contain demographic information.

## Conclusion

It is evident that the COVID-19 pandemic has inextricably changed walking behavior in US cities. Such large-scale alterations in walking patterns were inconceivable pre-COVID-19 pandemic. However, most importantly, our findings show that inequalities in walking opportunities among communities and neighborhoods have been reinforced, and new inequalities due to the interaction with COVID-19 response measures emerged. The experience of COVID-19 is not shared equally across our communities and neighborhoods, nor is its impact. As Marmot and Allen^25^ stated “COVID-19 exposes the fault lines in society and amplifies inequalities”. Our findings highlight walking inequality as an important indicator of disparities in the population and identify ‘walking poor’ subgroups, who importantly are most at risk from COVID-19. These subpopulations are set to benefit most from interventions and policies to promote walking and other physical activities. The novel methods applied in this work and our findings can help us to understand the prevalence, spread, and effects of walking within and across cities, countries and subgroups and to design communities, policies, and actions that promote greater walking in a COVID-19 secure world.

## Methods

### Mobility Data

The mobility data was obtained from Cuebiq, a location intelligence and measurement company. The dataset consists of anonymized records of GPS locations from anonymous users that opted-in to share the data anonymously in 10 metropolitan areas over a period of 5 months, from February to June 2020. Data was shared under a strict contract with Cuebiq through their Data for Good COVID19 Collaborative program where they provide access to de-identified and privacy-enhanced mobility data for academic research and humanitarian initiatives only. All researchers were contractually obligated to not share data further or to attempt to re-identify data. Mobility data is derived from anonymous users who opted in to share their data anonymously through a General Data Protection Regulation (GDPR) and California Consumer Privacy Act (CCPA) compliant framework.

Our sample dataset achieves broad geographic representation. Although the population and number of anonymous devices detected in the real data by census tract area is highly correlated (Pearson’s correlation of 0.66), post-stratification techniques were implemented to assure the representativeness of the data at the level of population^26^.

### Other data

Demographic data (median income, fraction of black people, fraction of users of public transportation, and fraction of people older than 64 yo) by census tract was obtained through the census ACS 5 year 2013-2018 datasets^27^. Obesity prevalence by census tract is given by the Center for Disease Control 500 cities estimations^28^. Park access was obtained from the City Health Dashboard^29^.

### Multivariate model

To study the combined effect of the demographic, physical environment, and health variables on walking behavior we use linear regression to explain 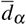, the average distance walked by people living in a census tract *α* as a function of those variables

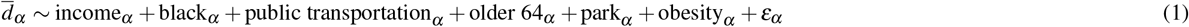

where *ε*_*α*_ is regression error. Results are presented in Table 1 where 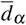 is calculated before and after COVID response measures (March 15th).

### Walk Detection

Walks were extracted from consecutive locations in which the speed between them did not exceed typical walking speeds (2 meters/second). Due to the geospatial accuracy of the data, we discarded walks of distance less than 50 meters.

### Walk classification

Walks have different purposes. The most common are recreational (leisure), transportation, shopping or going to work. Although in some cases it might be difficult to separate them (a recreational walk may involve some shopping), most of the literature separates walks by distance and/or destination. Leisure and utilitarian walks have very different lengths^8^ and, of course, most of the leisure walks happen in outdoor spaces or close to walkers’ residence.

In our data (see Figure 4) during weekdays, most of the walks occurred around 7am and from 3pm to 5pm. These walks were very short in length (below 750m) and are probably related to rush hour (i.e. travels to transportation)^8^, which is known to make up a significant proportion of walking behavior in US cities. Further evidence is provided by the disappearance of those clusters of walks during the weekends, being replaced by a cluster of very short walks around lunch time (noon). Also, after COVID-19 response measures were introduced, these patterns disappeared and most of the walks happened from 4pm to 6pm and with distances further than 750m. These findings suggest that there is a difference between walks below 750m which likely correspond mostly to utilitarian walking.

**Figure 4.**
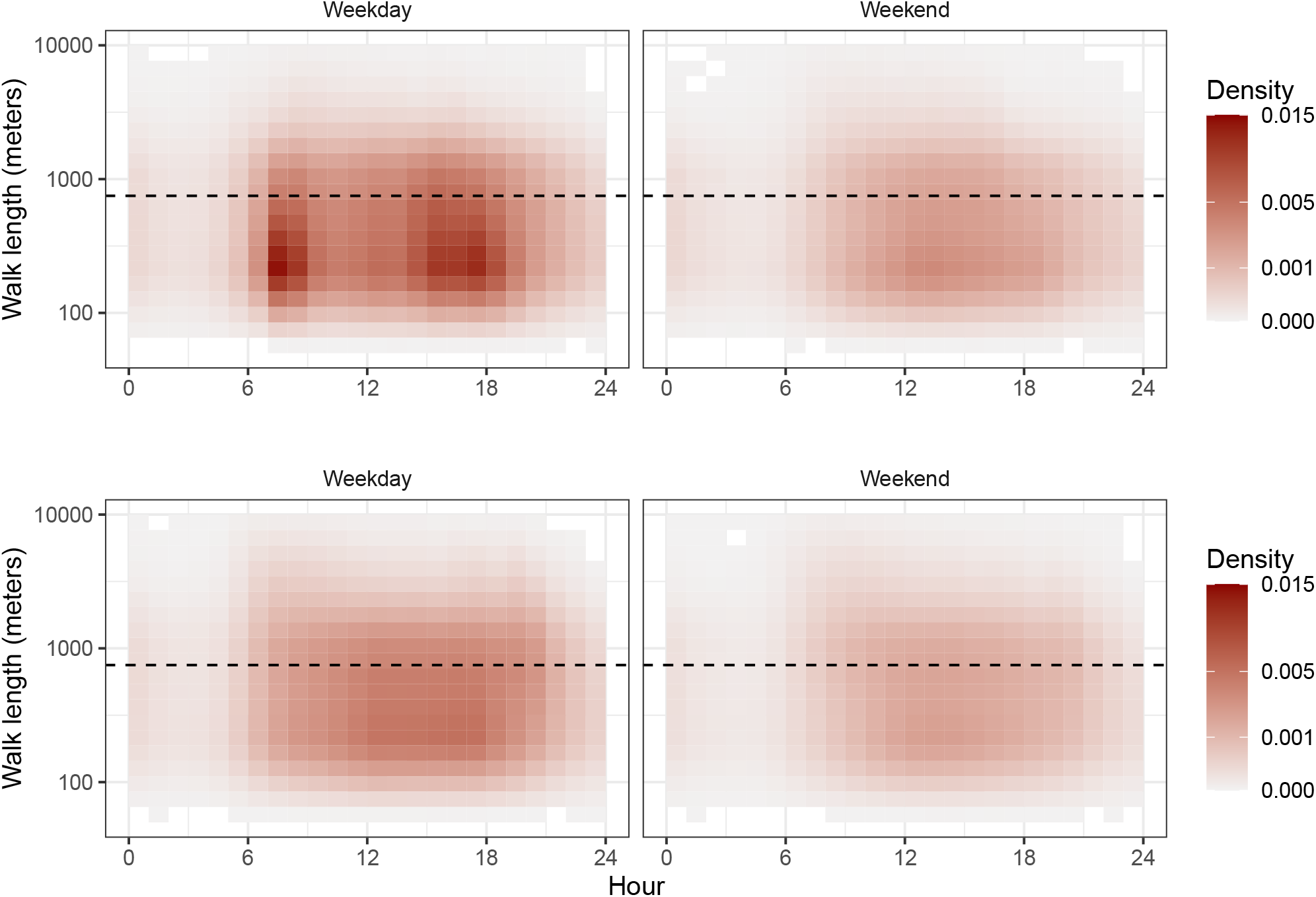
Panels A) to D) show the distribution of walks by hour of the day and length (in meters) for the walks before (upper panels) and after (lower panels) COVID-19 measures and for weekdays and weekends. Dashed line correspond to 750 meters.

On the other hand, walks have very different destinations. Even a walk longer than 750 meters could end up in a shopping, transportation or work venue. Using a large database of 1.5 Million Points of Interest (POI) from Foursquare we investigate the destination of the walks using the closest (up to 25 meters) POI to the last point of the walk before COVID-19 response measures. Table 3 shows the properties of the walks by different types of destination. In all the cities, round 74.71% of the walks do not end up in a particular POI of our dataset, while for the rest the most frequent destination is Food (5.39%), Service (4.94%), Shopping (3.01%) or Transportation (2.72%) categories. Shorter walks are to Shopping (453.95m), Grocery (489.31 meters) or Coffee / Tea (518.35 meters) while those without destination (708.94 meters) and City / Outdoors (639.01 meters) or Sports (617 meters) destinations are larger. Because of this variability for those walks longer than 750 meters we classify them as leisure if they do not end up on a particular POI of our dataset or if they end up in City / Outdoors (which includes residential areas).

**Table 3.**
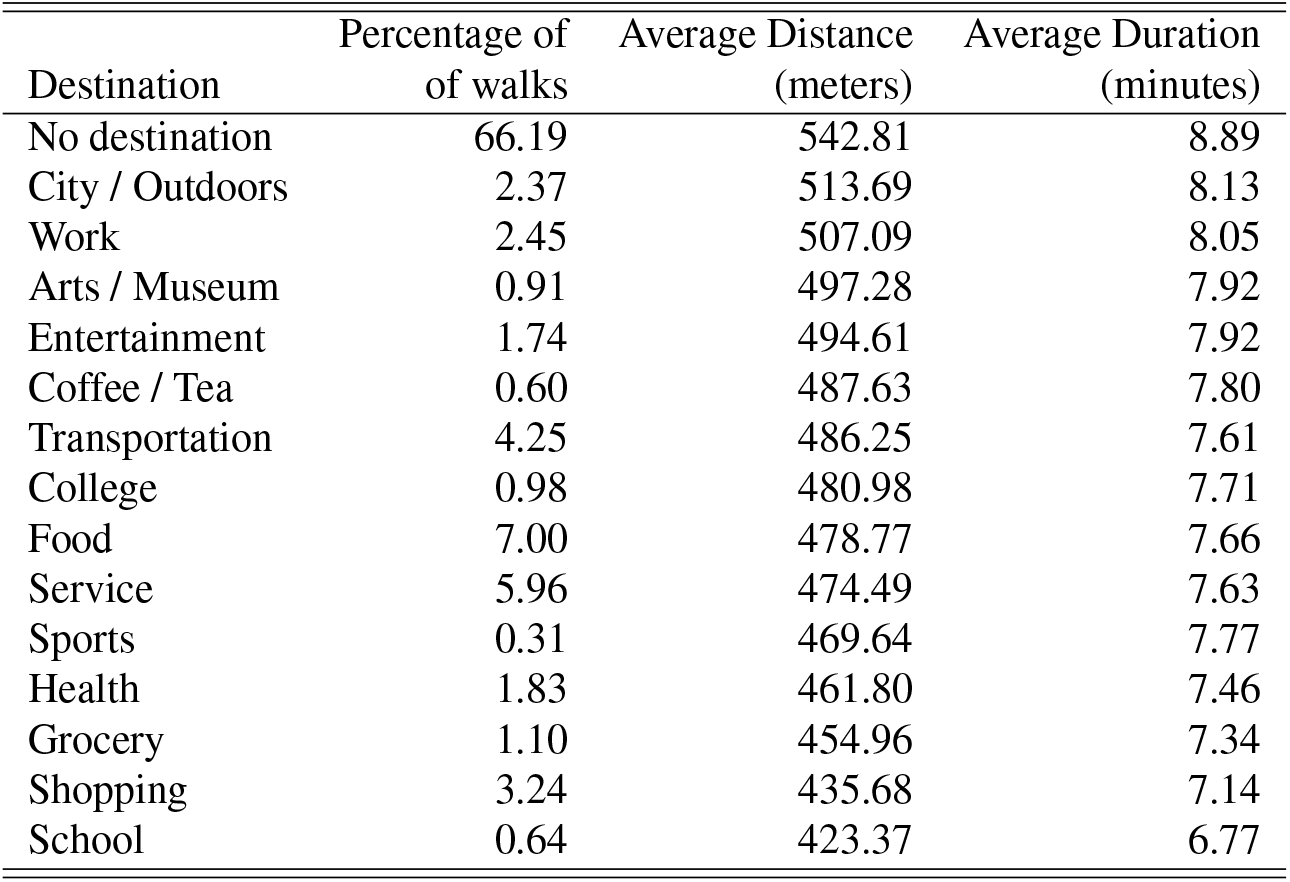
Percentage and properties of walks by destination. Ordered by average distance.

### Robustness of walk classification

Our results are robust regarding our criteria to define utilitarian walks. In particular, the large drop in the number of utilitarian walks after COVID-19 response measures were introduced and the fact that leisure walks have not decreased significantly is also observed for other distance thresholds (see Figure 5).

**Figure 5.**
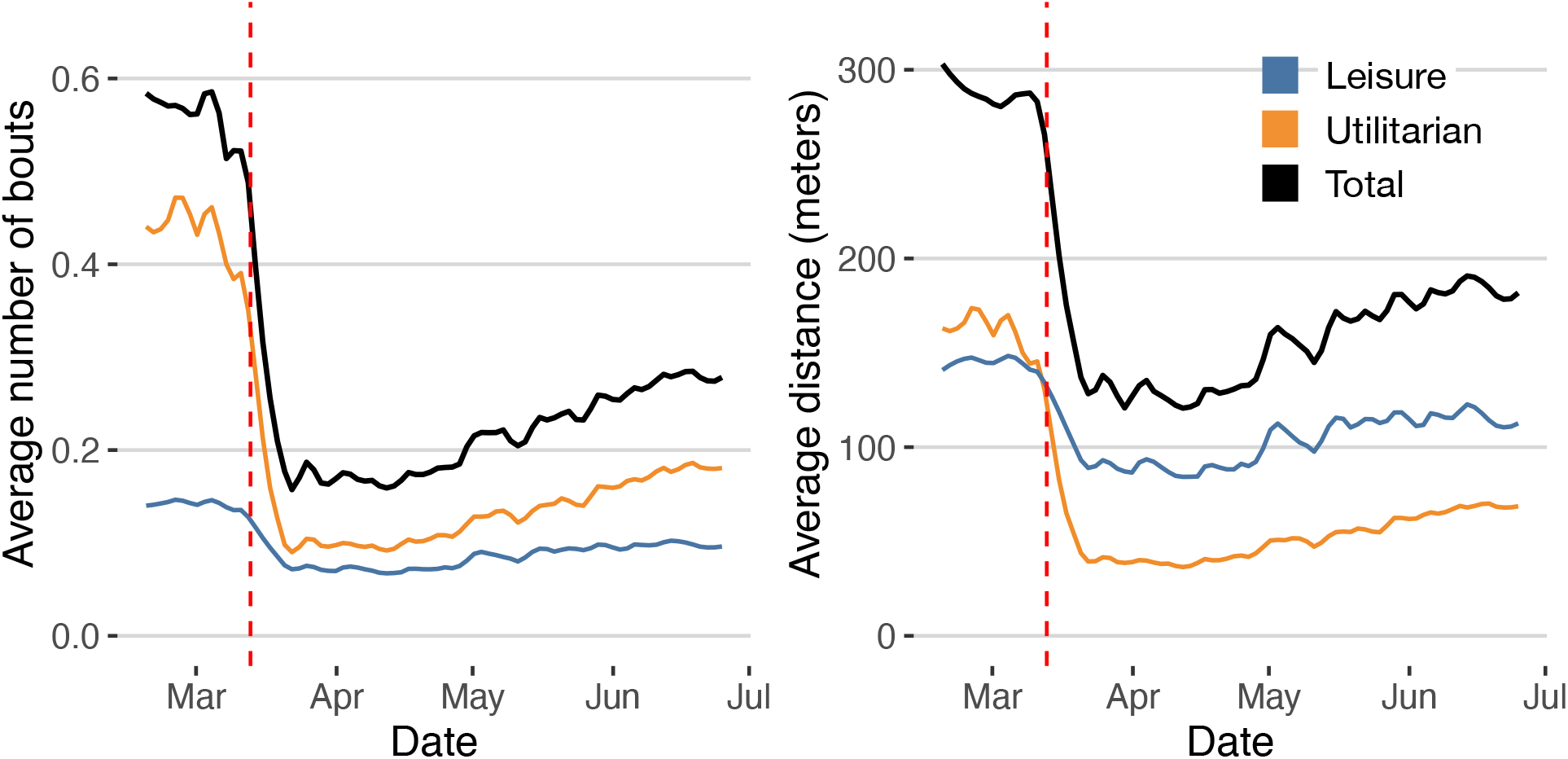
Time series of the average number of walks (left) and distance (right) by user and day using 500 meters as the threshold for utilitarian walks.

## Data Availability

The data that support the findings of this study is available from Cuebiq through their COVID-19 Collaborative program, but restrictions apply to the availability of these data, which were used under licenses for the current study, and so are not publicly available. Aggregated data used in the models are however available from the authors upon reasonable request and permission of Cuebiq.
Other data used comes from the American Community Survey (5-year) from the Census.

## Acknowledgements

We would like to thank Cuebiq for allowing the use of anonymized data through their COVID19 Collaborative program. EM acknowledges partial support by MINECO (FIS2016-78904-C3-3-P and PID2019-106811GB-C32). The funders had no role in study design, data collection, and analysis, decision to publish, or preparation of the manuscript.

## Author contributions statement

R.H, L.G, A.P and E.M designed and performed the research. E.M. Analyzed the data. All the authors wrote the paper.

## Additional information

The authors declare no competing interest.

To whom correspondence should be addressed: E.M.

